# Effects of photobiomodulation therapy combined with static magnetic field (PBMT-sMF) in patients with severe COVID-19 requiring intubation: a pragmatic randomized placebo-controlled trial

**DOI:** 10.1101/2020.12.02.20237974

**Authors:** Thiago De Marchi, Fabiano Frâncio, João Vitor Ferlito, Renata Monteiro Weigert, Cristiane Aparecida de Oliveira, Ana Paula Merlo, Délcio Luis Pandini, Bolivar Antônio Pasqual Júnior, Daniela Frare Giovanella, Shaiane Silva Tomazoni, Ernesto Cesar Pinto Leal-Junior

## Abstract

**Background:** Photobiomodulation therapy (PBMT) when used isolated or combined with static magnetic field (PBMT-sMF) has been proven benefits on skeletal muscle increasing performance and reducing fatigue, increasing oxygen saturation, and modulating inflammatory process. However, it is unknown whether the effects observed with this therapy on respiratory muscles will be similar to the effects previously observed on skeletal muscles.

**Objective:** We aimed to investigate whether PBMT-sMF is able to decrease the length of stay in the intensive care unit (ICU) and to reduce the mortality rate of patients with severe COVID-19 requiring invasive mechanical ventilation, increasing the respiratory function and modulating the inflammatory process.

**Methods:** We conducted a prospectively registered, pragmatic, triple-blinded (patients, therapists and outcome assessors), randomized, placebo-controlled trial of PBMT-sMF in patients with severe COVID-19, requiring invasive mechanical ventilation, admitted to the ICU. Patients were randomly assigned to receive either PBMT-sMF (6 sites at the lower thorax – 189 J total, and 2 sites at the neck area – 63 J total) or placebo PBMT-sMF daily during all the ICU stay. The primary outcome was length of stay in the ICU defined by either discharge or death. The secondary outcomes were survival rate, muscle function of diaphragm, change in blood tests, change in mechanical ventilation parameters and change in arterial blood gas analysis.

**Results:** A total of 30 patients underwent randomization (with 15 assigned to PBMT-sMF and 15 to placebo) and were analyzed. The length of stay in the ICU for the placebo group was 23.06 days while for the PBMT-sMF group was 16.26. However, there was no statistically difference between groups for the length of stay in the ICU (mean difference - MD = - 6.80; 95% CI = - 18.71 to 5.11). Regarding the secondary outcomes were observed statistically differences in favor of PBMT-sMF for diaphragm thickness, fraction of inspired oxygen, partial pressure of oxygen/fraction of inspired oxygen ratio, C-reactive protein, lymphocytes count, and hemoglobin (p<0.05).

**Conclusion:** Among patients with severe COVID-19 requiring invasive mechanical ventilation, PBMT-sMF was not statistically different than placebo to the length of stay in the ICU. However, it is important to highlight that our sample size was underpowered to detect statistical differences to the primary outcome. In contrast, PBMT-sMF increased muscle function of diaphragm, improved ventilatory parameters, decreased C-reactive protein levels and hemoglobin count, and increased lymphocytes count.

## INTRODUCTION

The novel Coronavirus disease 2019 (COVID-19) is caused by severe acute respiratory syndrome coronavirus 2 (SARS-CoV-2).^1^ SARS-CoV-2 infection trigger some blood changes such as leucopenia, lymphopenia, increased prothrombin time, d-dimer and c-reactive protein (PCR) levels, and increased cytokine levels such as 1L-1β and TNF-α.^2,3,4^ The main clinical features observed in patients with COVID-19 are fever, dry cough, and fatigue or myalgia;^2,5^ and can range from no symptoms to severe pneumonia and death.^1^ Patients with high degree of severity usually may progress to complications such as dyspnea, hypoxia, acute hypoxaemic respiratory insufficiency, arrhythmia, acute cardiac injury, and shock.^2,5,6^ These complications may require critical care in an intensive care unit (ICU).^2,5,7^

To date, there is no effective treatment against SARS-CoV-2 infection; therefore, several therapeutic agents such as chloroquine, hydroxychloroquine,^8^ lopinavir-ritonavir,^9^ remdesevir,^10^ and dexamethasone,^11^ often used to treat other medical conditions, have been tested and used in an attempt to face the COVID-19. The effectiveness of these therapeutic agents is still conflicting ^8-10^ and further high quality randomized controlled trials are necessary to confirm whether the benefits outweigh the harms. However, since there is no robust evidence about the effects of the available therapeutic agents and there is no effective treatment available to combat the SARS-CoV-2, it has been necessary to use management strategies of signs and symptoms in patients with COVID-19, especially in the most severe cases. The respiratory management of patients with severe COVID-19 in ICUs can be done through oxygen therapy, non-invasive ventilation and intubation.^12^ Respiratory failure due to hypoxemia is one of the most prominent complications in these patients^13^ and usually requires mechanical ventilation via an endotracheal tube.^14^ Acute respiratory failure reduces lung compliance, increases respiratory work and alters blood oxygenation, leading to a shallow breathing pattern.^15^ In addition, the strength of the respiratory muscles may also be reduced,^16^ which makes it difficult to successfully wean patients from mechanical ventilation, besides to contributing to a poorer clinical trajectory.^17,18^

Photobiomodulation therapy (PBMT) combined with static magnetic field (sMF) has potential to be a promising non-pharmacological tool in the respiratory management of patients with severe COVID-19. PBMT is a non-thermal and non-ionizing light therapy applied in the form of light amplification by the stimulated emission of radiation (LASER), light-emitting diodes (LEDs), and/or broadband irradiation in the visible and infrared spectra.^19^ PBMT increases cellular metabolism^20,21^ and microcirculation,^22^ oxygen availability,^23,24^ redox metabolism^25^ and modulates the inflammatory process.^26-28^ PBMT has been used in combination with a static magnetic field (sMF),^27-30^ generating better effects on cell metabolism.^31^ Robust evidence has shown that PBMT isolated or combined with sMF (PBMT-sMF) has beneficial effects on skeletal muscle, increasing performance and reducing fatigue,^32,33^ decreasing performance loss and function in detraining period,^30^ and increasing oxygen saturation.^23,24^ In contrast, there is a lack of evidence about the effects of PBMT or PBMT-sMF on respiratory muscles and respiratory system in general. To date, there is only one clinical trial that irradiated PBMT in respiratory muscles of the thorax and neck area.^34^ This study showed the effectiveness of PBMT in improving the functional capacity of patients with chronic obstructive pulmonary disease.^34^ Furthermore, experimental studies on respiratory system of animals observed that PBMT is able to modulate the pulmonary inflammation^35-37^ and relieve bronchial hyperresponsiveness.^37^

The beneficial effects previously showed added to the lack of adverse effects known to date, suggest that PBMT-sMF could be a safer alternative to the use of drugs in the treatment of patients with severe COVID-19. However, it is unknown whether the effects observed with PBMT (isolated or PBMT-sMF) on respiratory muscles will be similar to the effects previously observed on skeletal muscles. In addition, it is unknown whether PBMT-sMF can modulate the inflammatory process in this particular disease and contribute to a clinical improvement of these patients. Thus, it is necessary to investigate whether PBMT-sMF is able to decrease muscle fatigue of respiratory muscles and loss of respiratory function, besides of increasing oxygen saturation, modulating the inflammatory process and, consequently, contributing to general improvement of patients with severe COVID-19. In addition, it is necessary to investigate whether these possible benefits of PBMT-sMF can contribute to accelerate the weaning process of mechanical ventilation and to decrease the length of stay in the ICU of patients with severe COVID-19. Therefore, we aimed to investigate whether PBMT-sMF is able to decrease the length of stay in the ICU and to reduce the mortality rate of patients with severe COVID-19 requiring invasive mechanical ventilation, increasing the respiratory function and modulating the inflammatory process.

## METHODS

### Trial design

A prospectively registered (NCT04386694), pragmatic, two-arms, parallel randomized, triple-blinded (patients, therapists and outcome assessors), placebo-controlled trial was conducted. This study adheres to CONSORT guidelines (Supplement 2) There were two deviations from the registered protocol. The first one was due our estimation that the endpoint assessment would be up to 20 days after randomization, because the patients would be discharged or dead from any cause within this period. However, as the endpoint assessment directly depended on the length of stay in the ICU, it could take more than 20 days. The second deviation was regarding the assessment of three secondary outcomes: immunoglobulin G (IgG), immunoglobulin M (IgM), and D-dimer. These secondary outcomes were not assessed because the third part laboratory in charge to carry out the blood analysis was not able to implement the necessary routines before the beginning of this trial.

### Ethics

This study was submitted and approved by the Research Ethics Committee of Associação Dr. Bartholomeu Tacchini/Hospital Tacchini/RS, under protocol number 3,985,226, and by the National Research Ethics Commission from Brazilian Ministry of Health (protocol number 4,021,485). All patients eligible for the study or patient’s legal representative (if the patient was too unwell to provide consent) were informed by study assessors of the objective and all signed the written informed consent before enrollment in the study. Research personnel were taken all appropriate and customary steps to ensure that data remained secure and that patient privacy and confidentiality was maintained.

### Participants and recruitment

Participants were patients with laboratory-confirmed COVID-19 through RT-PCR (reverse transcription-polymerase chain reaction), admitted to the adult ICU of the Hospital Tachhini, Bento Gonçalves, Brazil, between May 2020 and July 2020. To be eligible, patients had to have 15 years or older; requiring invasive mechanical ventilation, through orotracheal intubation, due to respiratory failure. Patients who had a negative result in the diagnostic examination for COVID-19 and patients in prone position for more than 24 hours were excluded. Moreover, cancer patients and pregnant women were also excluded.

### Randomization and blinding

Prior to initiation of the treatment, patients were randomized into their respective intervention groups: active PBMT-sMF or placebo PBMT-sMF. The randomization was generated by a website (http://randomization.com/) and performed by a participating researcher not involved with the recruitment, assessment or treatment of patients. This same researcher was responsible for programming the PBMT-sMF device according to the result of the randomization, as active or placebo mode. This researcher was instructed not to disclose the programmed intervention to the assessor, therapist or any of the patients and other researchers involved in the study until its completion. The assessors, patients and therapists were blinded throughout the treatment. Concealed allocation was achieved through the use of sequentially numbered, sealed and opaque envelopes.

### Interventions

Patients were randomly allocated to two groups to be submitted to the active PBMT-sMF or placebo PBMT-sMF interventions. All patients, regardless of the allocated group, received standard ICU care associated with the tested intervention (active or placebo PBMT-sMF). The active and placebo PBMT-sMF were performed using the same device and the irradiated sites were the same to both therapies (Figure 1). To ensure blinding for therapists the device emitted the same sounds and the same information on the display regardless of the programmed mode (active or placebo). Furthermore, since this technology produces an undiscernible amount of heat,^29^ the blindness was not compromised by this aspect.

**Figure 1.**
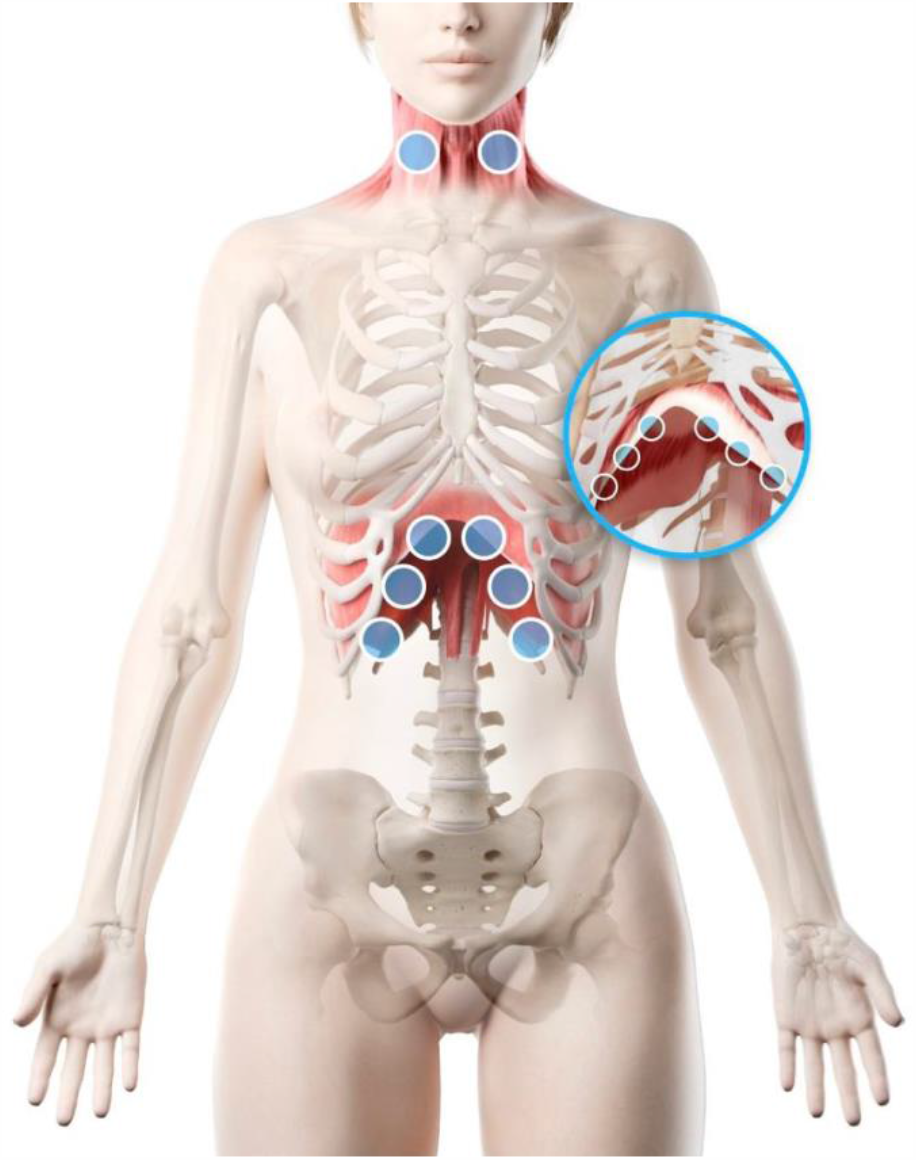
Sites of PBMT-sMF irradiation.

Patients underwent the treatment (active PBMT-sMF or placebo PBMT-sMF) according to prior randomization, once a day, during all the ICU stay (after inclusion in the study), until discharge or death. Specifications of the intervention:

1) Photobiomodulation therapy (PBMT) combined with static magnetic field (sMF) (PBMT-sMF) - active: PBMT-sMF was performed employing a cordless, portable MR5 Activ Pro LaserShower™ device (PhotOxyl™ prototype, manufactured by Multi Radiance Medical™, Solon - OH, USA). PBMT-sMF was irradiated in six sites at the lower thorax/ upper abdominal cavity and two sites at the neck area (sternocleidomastoid muscle), as illustrated in figure 1. PBMT-sMF exposure time was 60 seconds per site. The dose used in the lower thorax was 31.50 J per site, totalizing a dose of 189 J. In addition, the dose used in the neck area was 31.50 J per site, totalizing a dose of 63 J. PBMT-sMF dose per site, and the irradiation sites were established based on the only one previous study that have irradiated respiratory muscles.^34^ PBMT-sMF was applied using the direct contact method with slight pressure on the skin. The full description of parameters is provided in table 1.

**Table 1.** PBMT-sMF parameters

|  | Diaphragm | Sternocleidomastoid /<br>Scalene |
| --- | --- | --- |
| Number of lasers | 4 | 4 |
| Wavelength (nm) | 905 | 905 |
| Frequency (Hz) | 250 | 250 |
| Peak power (W) - each | 50 | 50 |
| Average mean optical output (mW) - each | 1.25 | 1.25 |
| Power density (mW/cm<sup>2</sup>) - each | 3.91 | 3.91 |
| Energy density (J/ cm<sup>2</sup>) - each | 0.234 | 0.234 |
| Dose (J) - each | 0.075 | 0.075 |
| Spot size of laser (cm<sup>2</sup>) - each | 0.32 | 0.32 |
| Number of red LEDs | 8 | 8 |
| Wavelength of red LEDs (nm) | 633 | 633 |
| Frequency (Hz) | 2 | 2 |
| Average optical output (mW) - each | 25 | 25 |
| Power density (mW/cm<sup>2</sup>) - each | 29.41 | 29.41 |
| Energy density (J/ cm<sup>2</sup>) - each | 1.765 | 1.765 |
| Dose (J) - each | 1.50 | 1.50 |
| Spot size of red LED (cm<sup>2</sup>) - each | 0.85 | 0.85 |
| Number of infrared LEDs | 8 | 8 |
| Wavelength of infrared LEDs (nm) | 850 | 850 |
| Frequency (Hz) | 250 | 250 |
| Average optical output (mW) - each | 40 | 40 |
| Power density (mW/cm<sup>2</sup>) - each | 71.23 | 71.23 |
| Energy density (J/ cm<sup>2</sup>) - each | 4.286 | 4.286 |
| Dose (J) - each | 2.40 | 2.40 |
| Spot size of infrared LED (cm<sup>2</sup>) - each | 0.56 | 0.56 |
| Magnetic field (mT) | 110 | 110 |
| Irradiation time per site (sec) | 60 | 60 |
| Total dose per site (J) | 31.50 | 31.50 |
| Number of irradiated sites | 6 | 1 (bilaterally) |
| Total dose delivered to the muscle group (J) | 189.00 | 31.50 (bilaterally) |
| Aperture of device (cm<sup>2</sup>) | 33 | 33 |
| Application mode | Cluster probe held stationary in skin contact with a 90-degree angle and slight pressure | Cluster probe held stationary in skin contact with a 90-degree angle and slight pressure |

2) Photobiomodulation therapy (PBMT) combined with static magnetic field (sMF) (PBMT-sMF) – placebo: The placebo intervention was delivered using the same device than active PBMT-sMF but without any emission of therapeutic dose. In the placebo mode, the infrared laser diodes, the infrared LED diodes, and the sMF were deactivated (turned off), and the power of the red LED diodes were decreased to 0.5 mW (mean power for each diode) in order to keep the visual aspect of red light, but not to deliver an effective therapeutic or considerable dose (0.24 J to each site) according the current available evidence.^19,33^ Moreover, the irradiated sites and the exposure time were the same that active PBMT-sMF.

### Outcomes

Demographic and clinical characteristics (e.g., age, gender, and comorbidities) were collected directly from electronic medical record of each patient. The primary and secondary outcomes were:

#### Primary outcome

- Length of stay in the ICU: the length of stay in the ICU was measured by the number of days that patients were maintained hospitalized in the ICU from randomization until discharge or death from any cause, whichever came first.

#### Secondary outcomes

- Survival rate: survival rate was measured by the rate of how many people survived and were discharged from ICU versus how many people have died, from randomization until discharge or death from any cause, whichever came first.
- Muscle function of diaphragm: the muscle function of the diaphragm was assessed through the thickness of diaphragm measured by ultrasound.^38,39^ The LOGIQe device (GE Healthcare, Chicago, USA) was used, with a linear transducer (ML6-15 from 5 to 15 MHz and 9L 2-8 MHz). The measurement was performed with the patient in the supine position. The transducer was positioned in the zone of apposition between the anterior and midaxillary lines at the level of the 9th or 10th intercostal space.^39^ End-expiratory thickness of diaphragm was measured in 2 consecutive breaths from 2 separate images. Measurements were repeated at least once until consistently within 10%; the mean of all 4 measurement was used for analysis.^39^ The measurement was obtained at baseline (up to 24 hours after the initiation of invasive mechanical ventilation), 10 days after randomization and in the last test before discharge or death from any cause, whichever came first (within 24 hours).
- Change in blood tests: the change in C-reactive protein (CRP), tumor necrosis factor-alpha (TNF-α), and vitamin D levels, besides change in erythrocytes, hemoglobin, hematocrit, leucocytes, segmented neutrals, eosinophiles, basophiles, lymphocytes, monocytes, and platelet count were assessed at baseline (admission to the ICU), 10 days after randomization and in the last test/day before discharge or death from any cause, whichever came first. The data regarding the blood tests were collected directly from electronic medical record of each patient, since it is a hospital daily routine to perform these blood tests. The data were collected by two assessors blinded to the allocation group of the patients.
- Change in mechanical ventilation parameters: the change in positive end-expiratory pressure levels (PEEP) and fraction of inspired oxygen (FiO_2_) were measured using a mechanical ventilator. The data was collected directly from the mechanical ventilator at baseline (admission to the ICU), 10 days after randomization and in the last day before discharge or death from any cause, whichever came first.
- Change in arterial blood gas analysis: the change in arterial partial pressure of oxygen (PO_2_) and PO_2_/FiO_2_ ratio were assessed at baseline (admission to the ICU), 10 days after randomization and in the last test/day before discharge or death from any cause, whichever came first. The data regarding the arterial blood gas analysis were collected directly from electronic medical record of each patient, since it is a hospital daily routine to perform these blood tests. The data were collected by two assessors blinded to the allocation group of the patients.

### Statistical analysis

Since to date there is not published studies aiming to assess the effects of PBMT-sMF in patients with severe COVID-19, we were compelled to use a convenience sample for this trial. To estimate our sample, we based on the number of patients that would fulfill our inclusion/exclusion criteria admitted in the ICU of Hospital Tacchini at the month before we start our trial (April 2020), which was 10 patients. Thus, estimating a 3-month length for the inclusion of patients in our trial (after the randomization of the first patient) we expected to reach the convenience sample of 30 patients in total.

The statistical analysis was conducted following intention-to-treat principles (i.e., the participants were analyzed in the groups to which they were allocated).^40,41^ Data normality was tested by Kolmogorov-Smirnov test. Since the data showed normal distribution, the between-group differences (effects of treatment) were analyzed by unpaired, two tailed, t tests (hospitalization data), and two-way repeated measures analysis of variance (time *vs* experimental group) with post hoc Bonferroni correction (ventilatory parameters, biochemical markers and hemogram parameters). The association between categorical variables was analyzed using the Chi-square test. Data were expressed as mean and standard deviation, mean difference between treatments, and 95% confidence intervals (95% CIs). Data were also expressed as frequency (%). The significance level was set at p<0.05. The magnitude of differences (Cohen-d) between groups, to examine practical significances, was calculated using the mean and SD of placebo and PBMT-sMF treatments (using Gpower 3.1). We adopted the criteria of Cohen for the analysis (0.2: small; 0.50: moderate; 0.80: large). All analyses were calculated by one of the researchers who was not involved in data collection.

## RESULTS

### Patients

Of the 62 patients who were assessed for eligibility, 30 were included in the study and underwent randomization; 15 were assigned to the placebo group and 15 were assigned to the PBMT-sMF. All patients received the treatment as assigned (figure 2). The mean age of the patients was 66.06 years, the mean height was 166.53 cm, the mean body mass was 75.18 kg, and 53.33% of patients were male. The demographic and clinical characteristics of patients at baseline were similar (p>0.05) in both groups and are described in Table 2. There were no adverse effects observed in both groups.

**Table 2.** Demographic and clinical characteristics of the patients at baseline

|  | **Placebo (n = 15)** | **PBMT-sMF (n = 15)** |
| --- | --- | --- |
| Age (years) | 67.93 (11.68) | 64.2 (18.67) |
| Gender (%) |  |  |
| Female | 8 | 6 |
| Male | 6 | 8 |
| Body mass (Kg) | 77.96 (18.49) | 72.40 (16.94) |
| Height (cm) | 165.06 (11.57) | 168 (7.98) |
| Comorbidities (%) |  |  |
| Hypertension | 9 (60.0) | 5 (33.3) |
| Diabetes | 8 (53.3) | 4 (26.7) |
| Obesity | 4 (26.7) | 3 (20.0) |
| Dementia | 3 (20.0) | 2 (13.3) |
| Depression | 2 (13.3) | 0 (0.00) |

Categorical variables are expressed as number (%). Continuous variables are expressed as mean (SD). PBMT-sMF: photobiomodulation therapy combined with static magnetic field.

**Figure 2.**
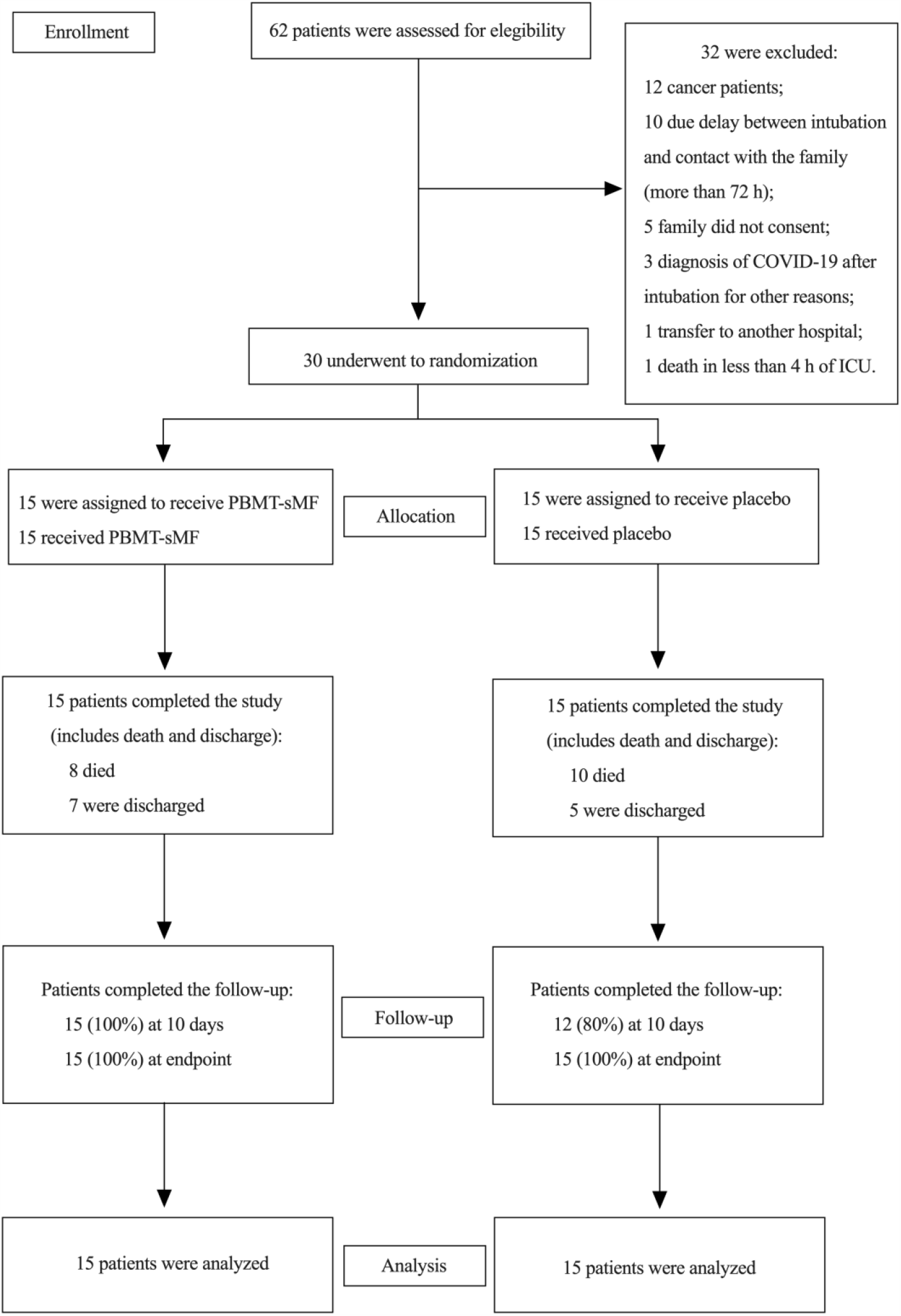
Enrollment and randomization.

### Primary outcome

The length of stay in the ICU for the placebo group was 23.06 days while for the PBMT-sMF group was 16.26 days. However, there was no statistically difference between groups for the length of stay in the ICU (mean difference – MD = - 6.80; 95% CI = - 18.71 to 5.11) (table 3).

**Table 3.** Hospitalization outcomes

|  | **Placebo** | **PBMT-sMF** | **Between groups comparisons** | **Mean difference [95% CI]** | **Effect size Cohen d** |
| --- | --- | --- | --- | --- | --- |
| Length of stay in ICU (Days) | Total (n=15): 23.06 (± 20.37)<br>Non-survivors (n=10): 20.4 (± 20.58)<br>Survivors (n=5): 28.4 (± 21.12) | Total (n=15): 16.26 (± 9.61)<br>Non-survivors (n=8): 18.65 (± 9.88)<br>Survivors (n=7): 13.57 (± 9.25) | Total: p=0.25<br>Non-survivors: p=0.82<br>Survivors: p=0.12 | Total: - 6.80 [-18.71 to 5.11]<br>Non-survivors: -1.75 [-18.6 to 15.1]<br>Survivors: -14.83 [-34.61 to 4.94] | Total: 0.4 (small)<br>Non-survivors: 0.1 (small)<br>Survivors: 0.9 (large) |
| Hospitalization length (Days) | Total (n=15): 24.33 (± 20.24)<br>Non-survivors (n=10): 20.40 (± 20.58)<br>Survivors (n=5): 32.20 (± 19.09) | Total (n=15): 18.33 (± 8.95)<br>Non-survivors (n=8): 18.75 (± 10.08)<br>Survivors (n=7): 17.85 (± 8.17) | Total: p=0.30<br>Non-survivors: p=0.83<br>Survivors: p=0.10 | Total: -6.0 [-17.7 to 5.69]<br>Non-survivors: -1.65 [-18.56 to 15.26]<br>Survivors: -14.35 [-32.13 to 3.43] | Total: 0.4 (small)<br>Non-survivors: 0.1 (small)<br>Survivors: 0.9 (large) |
| Deaths / Discharges | 10 / 5 | 8 / 7 | p=0.46 | ----- | ----- |

Continuous variables are expressed as mean (SD). Categorical variables are expressed as number. PBMT-sMF: photobiomodulation therapy combined with static magnetic field; ICU: intensive care unit.

### Secondary outcomes

Patients allocated to the PBMT-sMF group increased diaphragm thickness at the assessment performed at day 10 (MD = 14.19; 95% CI = 2.50 to 25.87) and at endpoint assessment (MD = 20.46; 95% CI = 8.77 to 32.14) compared to patients allocated to the placebo group (table 4). Regarding the ventilatory parameters, there was no statistically difference between groups for PEEP and PO_2_ at any time point (table 4). In addition, there was no statistically difference between groups for FiO_2_ at assessment of day 10. However, PBMT-sMF group was able to decrease the FiO_2_ at endpoint assessment (MD = - 23.93; 95% CI = - 47.52 to - 0.34) compared to placebo group (table 4). Finally, patients allocated to the PBMT-sMF group increased PO_2_/FiO_2_ only at endpoint assessment (MD = 117.90; 95% CI =12.31 to 223.50) compared to the placebo group (table 4).

**Table 4.** Ventilatory parameters

|  | Placebo (n = 15) |  |  | PBMT-sMF (n = 15) |  |  | Placebo vs PBMT-sMF |  |  |
| --- | --- | --- | --- | --- | --- | --- | --- | --- | --- |
|  | Baseline | Day 10 | Endpoint | Baseline | Day 10 | Endpoint | Between groups comparisons | Mean difference [95% CI] | Effect size Cohen d |
| Thickness fraction diaphragm (%) | 25.28<br>(± 12.85) | 22.90<br>(± 7.58) | 23.76<br>(± 13.15) | 29.55<br>(± 13.56) | 37.09<br>(± 17.02) | 44.22<br>(± 12.59)** | Baseline: p=0.99<br>Day 10: p=0.0117<br>Endpoint: p=0.0001 | Baseline: 4.27<br>[-7.41 to 15.95]<br>Day 10: 14.19<br>[2.50 to 25.87]<br>Endpoint: 20.46<br>[8.77 to 32.14] | Day 10: 1.1<br>(large)<br>Endpoint: 1.6<br>(large) |
| PEEP (cmH<sub>2</sub>O) | 9.13<br>(± 2.64) | 8.93<br>(± 2.46) | 8.20<br>(± 4.09) | 9.00<br>(± 1.96) | 7.53<br>(± 2.82) | 6.60<br>(± 2.82) | Baseline: p=0.99<br>Day 10: p=0.5540<br>Endpoint: p=0.3931 | Baseline: -0.13<br>[-2.69 to 2.43]<br>Day 10: -1.40<br>[-3.96 to 1.16]<br>Endpoint: -1.60<br>[- 4.16 to 0.96] | Day 10: 0.5<br>(moderate)<br>Endpoint: 0.5<br>(moderate) |
| PO<sub>2</sub> (mmHg) | 91.46<br>(± 20.78) | 99.74<br>(± 29.93) | 92.74<br>(± 28.00) | 108.22<br>(± 29.68) | 106.46<br>(± 33.98) | 109.29<br>(± 26.16) | Baseline: p=0.99<br>Day 10: p=0.99<br>Endpoint: p=0.34 | Baseline: 16.76<br>[-8.64 to 42.16]<br>Day 10: 6.72<br>[-18.68 to 32.12]<br>Endpoint: 16.55<br>[-8.85 to 41.95] | Day 10: 0.2<br>(small)<br>Endpoint: 0.6<br>(moderate) |
| FiO<sub>2</sub> (%) | 69.66<br>(± 21.91) | 63.20<br>(± 26.72) | 75.46<br>(± 31.59) | 62.33<br>(± 21.03) | 51.26<br>(± 26.07) | 51.53<br>(± 29.68) | Baseline: p= 0.33<br>Day 10: p=0.6590<br>Endpoint: p=0.0456 | Baseline: -7.33<br>[-30.92 to 16.26]<br>Day 10: -11.94<br>[- 35.53 to 11.65]<br>Endpoint:- 23.93<br>[-47.52 to -0.34] | Day 10: 0.5<br>(moderate)<br>Endpoint: 0.8<br>(large) |
| PO<sub>2</sub> / FiO<sub>2</sub> (mmHg) | 146.71<br>(± 61.85) | 183.32<br>(± 81.85) | 158.26<br>(± 102.37) | 199.65<br>(± 109.82) | 261.28<br>(± 156.21) | 276.15<br>(± 163.07) | Baseline: p=0.67<br>Day 10: p=0.2246<br>Endpoint: p=0.0233 | Baseline: 52.94<br>[-53.63 to 158.50]<br>Day 10: 77.95<br>[-27.63 to 183.5]<br>Endpoint: 117.90<br>[12.31 to 223.50] | Day 10: 0.6<br>(moderate)<br>Endpoint: 0.9<br>(large) |

\*\* Intragroup difference (PBMT-sMF) compared to the baseline (p<0.01). Continuous variables are expressed as mean (SD). PBMT-sMF: photobiomodulation therapy combined with static magnetic field.

Patients allocated to the PBMT-sMF group decreased the CRP levels (MD = - 83.87; 95% CI = - 166.50 to - 1.28) and increased the lymphocytes count (MD = 1160.00; 95% CI = 197.90 to 2122.00) only at endpoint assessment compared to placebo group (table 5). In addition, patients allocated to the PBMT-sMF group decreased the hemoglobin count (MD = - 1.97; 95% CI =- 3.77 to - 0.17) at assessment of day 10 compared to placebo group (table 5). Finally, there were no statistically significant differences between the groups for any other secondary outcomes at any time point.

**Table 5.** Biochemical markers and hemogram parameters

|  | Placebo (n = 15) |  |  | PBMT-sMF (n = 15) |  |  | Between groups comparisons | Mean difference [95% CI] | Effect size Cohen d |
| --- | --- | --- | --- | --- | --- | --- | --- | --- | --- |
|  | Baseline | Day 10 | Endpoint | Baseline | Day 10 | Endpoint |  |  |  |
| CRP (mg/dL) | 201.30<br>(± 65.39) | 118.79<br>(± 71.00) | 156.53<br>(± 124.49) | 152.29<br>(± 108.88) | 98.70<br>(± 95.24) | 72.66<br>(± 75.40) | Baseline:<br>p=0.41<br>Day 10:<br>p=0.99<br>Endpoint:<br>p=0.045 | Baseline: - 49.01<br>[-131.60 to 33.58]<br>Day 10: -20.09<br>[-102.70 to 62.50]<br>Endpoint: -83.87<br>[-166.50 to -1.28] | Day 10: 0.2<br>(small)<br>Endpoint: 0.8<br>(large) |
| TNF-alpha (pg/mL) | 17.23<br>(± 13.46) | 12.28<br>(± 5.42) | 17.05<br>(± 7.54) | 19.71<br>(± 17.51) | 18.40<br>(± 13.43) | 14.19<br>(± 5.44) | Baseline:<br>p=0.99<br>Day 10:<br>p=0.44<br>Endpoint:<br>p=0.99 | Baseline: 2.48<br>[-7.71 to 12.68]<br>Day 10: 6.11<br>[-4.08 to 16.32]<br>Endpoint: - 2.87<br>[-13.07 to 7.33] | Day 10: 0.5<br>(moderate)<br>Endpoint:0.4<br>(small) |
| Vitamin D (ng/mL) | 14.88<br>(± 5.13) | 18.28<br>(± 9.95) | 14.16<br>(± 4.80) | 19.04<br>(± 10.83) | 17.85<br>(± 6.67) | 21.07<br>(± 7.07) | Baseline:<br>p=0.43<br>Day 10:<br>p=0.99<br>Endpoint:<br>p=0.0502 | Baseline: 4.15<br>[-2.75 to 11.07]<br>Day 10: -0.42<br>[-7.34 to 6.48]<br>Endpoint: 6.90<br>[-0.004 to 13.82] | Day 10: 0.1<br>(small)<br>Endpoint:1.1<br>(large) |
| Erythrocytes (millions/mm<sup>3</sup>) | 4.20<br>(± 0.58) | 3.73<br>(± 0.55) | 3.40<br>(± 0.54)<sup>##</sup> | 3.75<br>(±0.64) | 3.18<br>(± 0.59) | 3.04<br>(± 0.90)<sup>**</sup> | Baseline:<br>p=0.19<br>Day 10:<br>p=0.065<br>Endpoint:<br>p=0.389 | Baseline: -0.45<br>[-1.02 to 0.13]<br>Day 10: -0.55<br>[-1.13 to 0.02]<br>Endpoint: -0.36<br>[-0.94 to 0.21] | Day 10: 0.9<br>(large)<br>Endpoint: 0.5<br>(moderate) |
| Hemoglobin (g/dL) | 12.57<br>(± 1.91) | 11.47<br>(± 1.78) | 9.97<br>(± 1.73)<sup>##</sup> | 11.34<br>(± 2.14) | 9.50<br>(± 1.55)<sup>*</sup> | 9.03<br>(± 2.75)<sup>**</sup> | Baseline:<br>p=0.29 | Baseline: -1.23<br>[-3.03 to 0.56]<br>Day 10: -1.97 | Day 10: 1.2<br>(large) |
|  |  |  |  |  |  |  | Day 10:<br>p=0.026<br>Endpoint:<br>p=0.6173 | [-3.77 to -0.17]<br>Endpoint: -0.94<br>[-2.74 to 0.86] | Endpoint: 0.4<br>(small) |
| Hematocrit (%) | 36.92<br>(± 4.94) | 32.66<br>(± 5.46) | 30.57<br>(± 4.58)<sup>##</sup> | 33.67<br>(± 5.04) | 29.03<br>(± 4.04) | 27.78<br>(± 7.20)<sup>**</sup> | Baseline:<br>p=0.29<br>Day 10:<br>p=0.194<br>Endpoint:<br>p=0.459 | Baseline: -3.25<br>[-7.98 to 1.48]<br>Day 10: -3.62<br>[-8.36 to 1.10]<br>Endpoint: -2.79<br>[-7.52 to 1.94] | Day 10: 0.8<br>(large)<br>Endpoint: 0.5<br>(moderate) |
| Leukocytes (mm<sup>3</sup>) | 11466.00<br>(± 6677.06) | 14221.92<br>(± 5231.44) | 17742.66<br>(± 11630.20) | 12175.33<br>(± 9330.37) | 23449.13<br>(± 33682.68) | 23186.66<br>(± 21612.65) | Baseline:<br>p=0.99<br>Day 10:<br>p=0.476<br>Endpoint:<br>p=0.99 | Baseline: 709.30<br>[-15147 to 16566]<br>Day 10: 9227<br>[-6629 to 25083]<br>Endpoint: 5444<br>[-10412 to 21300] | Day 10: 0.4<br>(small)<br>Endpoint: 0.3<br>(small) |
| Segmented Neutrals (mm<sup>3</sup>) | 9890.16<br>(± 5904.32) | 13010.82<br>(± 3191.26) | 13655.49<br>(± 10037.05) | 9834.14<br>(± 7425.83) | 10154.81<br>(± 3450.12) | 15017.58<br>(± 11566.73) | Baseline:<br>p=0.99<br>Day 10:<br>p=0.91<br>Endpoint:<br>p=0.99 | Baseline: -56.02<br>[-6836 to 6724]<br>Day 10: -2856<br>[-9636 to 3924]<br>Endpoint: 1362<br>[-5418 to 8142] | Day 10: 0.9<br>(large)<br>Endpoint: 0.1<br>(small) |
| Eosinophiles (mm<sup>3</sup>) | 47.43<br>(± 90.35) | 19.41<br>(± 29.58) | 1070.21<br>(± 2825.97)<sup>#</sup> | 49.73<br>(± 108.99) | 115.11<br>(± 114.62) | 223.86<br>(± 229.56) | Baseline:<br>p=0.99<br>Day 10:<br>p=0.99<br>Endpoint:<br>p=0.146 | Baseline: 2.30<br>[-1032 to 1037]<br>Day 10: 95.70<br>[-939 to 1130]<br>Endpoint:<br>- 846.40<br>[-1881 to 188.3] | Day 10: 1.1<br>(large)<br>Endpoint: 0.4<br>(small) |
| Basophiles (mm<sup>3</sup>) | 5.84<br>(± 6.07) | 7.83<br>(± 11.16) | 18.54<br>(± 27.53) | 7.57<br>(± 11.23) | 10.77<br>(± 18.75) | 22.88<br>(± 30.30) | Baseline:<br>p=0.99<br>Day 10:<br>p=0.99<br>Endpoint:<br>p=0.99 | Baseline: 1.73<br>[-15.79 to 19.26]<br>Day 10: 2.93<br>[-14.59 to 20.46]<br>Endpoint: 4.33<br>[-13.19 to 21.86] | Day 10: 0.2<br>(small)<br>Endpoint: 0.2<br>(small) |
| Lymphocyte (mm<sup>3</sup>) | 764.31<br>(± 415.39) | 1138.70<br>(± 484.81) | 1554.00<br>(± 1072.52) | 1183.26<br>(± 1224.40) | 1309.53<br>(± 560.02) | 2713.83 | Baseline:<br>p=0.87 | Baseline: 418.9<br>[-543 to 1381]<br>Day 10: 170.8 | Day 10: 0.3<br>(small) |
| | | | | | | (± 1899.26)**,<br>\$* | Day 10:<br>p=0.99<br>Endpoint:<br>p=0.0125 | [-791.1 to 1133]<br>Endpoint:<br>1160.00<br>[197.90 to 2122.00] | Endpoint: 0.8<br>(large) |
| Monocytes (mm<sup>3</sup>) | 557.06<br>(± 459.19) | 936.71<br>(± 499.14) | 1057.01 (± 646.43) | 661.23<br>(± 542.04) | 1224.89 (± 496.07)* | 1346.20<br>(± 862.56)** | Baseline:<br>p=0.99<br>Day 10:<br>p=0.576<br>Endpoint:<br>p=0.571 | Baseline: 104.2<br>[-431.3 to 639.6]<br>Day 10: 288.2<br>[-247.2 to 823.6]<br>Endpoint: 289.2<br>[-246.2 to 824.6] | Day 10: 0.6<br>(moderate)<br>Endpoint: 0.4<br>(small) |
| Platelets (mm<sup>3</sup>) | 189800.00<br>(± 92499.57) | 237890.86<br>(± 71588.08) | 231266.66<br>(± 104779.67) | 210028.26<br>(± 90254.29) | 261858.03<br>(± 145159.59) | 288800.00<br>(± 166270.09) | Baseline:<br>p=0.99<br>Day 10:<br>p=0.99<br>Endpoint:<br>p=0.54 | Baseline: 20228<br>[-83748 to 124204]<br>Day 10: 23967<br>[-80009 to 127943]<br>Endpoint: 57533<br>[-46443 to 161509] | Day 10: 0.2<br>(small)<br>Endpoint: 0.4<br>(small) |

\* Intragroup difference (PBMT-sMF) compared to the baseline (p<0.05); \$\* Intragroup difference (PBMT-sMF) compared to the Day 10 (p<0.01); \*\* Intragroup difference (PBMT-sMF) compared to the baseline (p<0.01); # Intragroup difference (Placebo) compared to the baseline (p<0.05); ## Intragroup difference (Placebo) compared to baseline (p<0.01). Continuous variables are expressed as mean (SD). PBMT-sMF: photobiomodulation therapy combined with static magnetic field

### Sample size calculation for further randomized controlled trials

Based on our results for the primary outcome (length of stay in the ICU), we are able to determine the sample size for further studies investigating the effects of PBMT-sMF in patients admitted to the ICU with COVID-19 requiring invasive mechanical ventilation.

It was observed that the length of stay in the ICU for patients treated with placebo was 23.06 (± 20.37), and for patients treated with PBMT-sMF it was 16.26 (± 9.61). Therefore, considering a β value of 20% and α of 5%, the calculation resulted in a sample of 68 patients per group. We used the SPH™ Analytics website to calculate the sample (https://www.sphanalytics.com/sample-size-calculator-using-average-values/).

## DISCUSSION

This is the first randomized triple-blinded placebo-controlled trial to assess the effects of PBMT-sMF in patients with severe COVID-19 requiring mechanical ventilation. We observed that length of stay in the ICU for PBMT-sMF group was shorter than to placebo group. However, there was no statistically difference between groups. In contrast, the use of PBMT-sMF increased diaphragm thickness at day 10 and at the endpoint assessment compared to placebo treatment. Furthermore, PBMT-sMF was superior to placebo in improving ventilatory parameters, decreasing FiO_2_ and increasing PO_2_/FiO_2_ at the endpoint assessment. In addition, the patients allocated to the PBMT-sMF group had decreased CRP levels and increased lymphocytes count at the endpoint assessment compared to the placebo group. Finally, PBMT-sMF was superior to placebo in the decrease of hemoglobin count at assessment of day 10. Regarding the other secondary outcomes, no between-group differences were observed.

Although there is speculation about the possible benefits of PBMT (isolated or combined with sMF) in patients with COVID-19,^42^ to date this is the only one randomized controlled trial carried out in this field. Therefore, the direct comparation with other randomized controlled trials is limited. However, we observed that the use of PBMT-sMF increased the muscle thickness and consequently its ability to generate strength,^43-45^ and also to prevent the harms due disuse^30^ as previously observed in skeletal muscles of humans. In addition, there is evidence that PBMT modulates the inflammatory process through decreased CRP levels also in skeletal muscle of humans,^46,47^ as we observed in our study. In regards of other treatments than PBMT to COVID-19, it has been shown that patients treated with lopinavir-ritonavir have a time to clinical improvement similar to the patients treated with standard care alone.^9^ In addition, treatment with convalescent plasma added to a standard treatment was similar to standard treatment alone regarding time to clinical improvement within 28 days.^48^ In contrast, patients that used remdesivir had a shorter time to recovery than patients treated with placebo.^10^ In our trial, we observed that patients treated with PBMT-sMF had shorter time to recovery as well. However, there was no difference between PBMT-sMF and placebo group, which may probably have been caused by the small sample size. It is important to highlight that we included only patients admitted to the ICU and requiring invasive mechanical ventilation, which is different than the aforementioned studies.^9,10,48^ Finally, we did not observe adverse effects with the use of PBMT-sMF, while the treatment with convalescent plasma added to a standard treatment, lopinavir-ritonavir, and remdesivir caused some adverse effects such as gastrointestinal events, chill and rashes, acute kidney injury and even serious respiratory failure.^9,10,48^

PBMT-sMF was able to improve some ventilatory parameters, besides inflammatory and infectious process and immune response in patients with severe COVID-19 requiring mechanical ventilation. It was previously observed that patients with severe COVID-19 present elevated ventilation-perfusion mismatch due to high dead space fraction.^49^ In our study, we observed a better ventilation/perfusion rate associated to a preservation and improvement of diaphragm thickness in patients treated with PBMT-sMF. These improvements possibly led to decreased inflammation and infection (CRP) helping the immune response (lymphocytes) of these patients. These findings suggest that the preservation of the main respiratory muscle may facilitate the process of weaning from mechanical ventilation and trigger a cascade of positive effects, improving the clinical condition of the patients. We observed that patients in the PBMT-sMF group had shorter length of stay in the hospital, considering both all patients and only the survivors. Therefore, these findings suggest that treatment with PBMT-sMF may reduce the burden caused in the hospital and health systems, and the use of scarce health care resources during this pandemic. In addition, PBMT-sMF has proven to be a safe therapy as there were no adverse effects with its use. In addition, the number of deaths in the PBMT-sMF group was smaller than in the placebo group, although there was no statistically difference between groups. This trial was prospectively registered, we used true randomization, concealment allocation, blindness of therapists, outcome assessors and patients. Statistical analysis was conducted following intention-to-treat principles and it was performed by a blind researcher for the treatment’ allocation. Moreover, we used a placebo group to control for confounders such as placebo effect, regression to mean, and natural recovery. However, although we have included several features in order to minimize bias, this study has some limitations. There were two deviation from the registered protocol. The first one was that we estimated that the endpoint assessment would be up to 20 days after randomization, however, as the endpoint assessment directly depended on the length of stay in the ICU, in some cases this assessment took more than 20 days. The second deviation was not having assessing IgG, IgM and D-dimmer because the third part laboratory in charge to carry out the blood analysis was not able to implement the necessary routines before the beginning of our trial. In addition, we might considerer as a limitation did not following up the patients after hospital discharge. Finally, another limitation to be considered is the small sample size.

This trial is the first one assessing the effects of PBMT-sMF in patients with severe COVID-19, therefore was unknown a priori the adequate sample size to detect precise differences in the primary outcome of the study. However, this study was important to estimate the adequate sample size for further studies considering length of stay in the ICU as a primary outcome. Based in our results we estimated that further studies will require *n*= 68 patients per group to detect clinically important difference between groups. Therefore, further studies with rigorous methodological quality and adequate sample size are needed to investigate whether PBMT-sMF is able to decrease the length of stay in the ICU for patients with severe COVID-19 requiring invasive mechanical ventilation.

### Conclusion

Among patients with severe COVID-19 requiring invasive mechanical ventilation, PBMT-sMF was not statistically different than placebo to the length of stay in the ICU. However, it is important to highlight that our sample size was underpowered to detect statistical differences to the primary outcome. In contrast, PBMT-sMF increased diaphragm thickness, decreased FiO_2_, increased PO_2_/FiO_2_, decreased CRP levels and hemoglobin count, and increased lymphocytes count.

## Data Availability

The datasets used and/or analyzed during the current study are available from the corresponding author on reasonable request.

## ACKNOWLEDGMENT

The authors thank the entire multidisciplinary team that worked in the ICU of Hospital Tacchini, the Tacchini Institute for Health Research, and Marcos Vinícius Ferlito and Tauani de Souza by the contribution with the research team.

